# The Lifestyle Factors of Physical Activity and Diet Balance associated with HPV Infection

**DOI:** 10.1101/2022.07.14.22277082

**Authors:** Yantao Li, Mengping Liu, Peng Huang, Zhongzhou Yang, Anli Wang

**Author notes:** These authors contributed equally to this work (Yantao Li, Mengping Liu).

## Abstract

**Background:** Human wellbeing has been linked with lifestyle factors such as physical activity, diet balance, sleep quality, depression, and anxiety. However, few studies illustrate the relationship between such lifestyle factors and HPV infection. In this study, we demonstrate that lifestyle factors might be crucial for reducing the burden of cervical cancer or HPV-related cancer.

**Participants and Methods:** Participants were recruited through a digital eHealth platform without vaccination from May 2020 to August 2021. Both lifestyle factors and cervicovaginal mucus (CVM) samples to test for HPV outcomes were collected from each participant. In addition, the eHealth platform recorded age and gynecological diseases, which were adjusted to apply for both univariable and multivariable logistic regression. Furthermore, lifestyle factors and HPV serotype were categorized as low, intermediate, and high risk in order to conduct stratification analysis. Finally, lifestyle factors were studied in connection with sole and multiple HPV serotypes.

**Results:** We recruited 149 HPV positive and 346 HPV negative through HPV detection. Physical activity and diet balance were significantly associated with HPV infection in lifestyle factors (P-values < 0.001) after adjusting for age and gynecological diseases. However, stratified analysis showed three factors were insignificant for HPV infection – namely, sleep quality, depression, and anxiety. Most HPV infections involved a sole HPV serotype (83%), and diet balance was the most significant difference between sole and multiple HPV infections.

**Conclusions:** Among lifestyle factors, low physical activity or low diet balance can significantly increase HPV infection. In particular, diet balance might be related to the number of HPV serotypes. Our results suggest that exercising and regulating one’s diet may reduce the burden of HPV-related cancer.

## Introduction

Lifestyle behavior factors such as physical activity, diet balance, sleep quality, depression and anxiety impact personal wellness [1]. The current prevalence of physical inactivity is estimated to be 23% [2, 3]. Hence, the World Health Organization has recommended a 10% increase in physical activity by 2020 [Refs]. Though epidemiological research has shown that a diet balance is associated with decreasing cancer mortality [4, 5], related randomized controlled trials (RCT) for diet balance have not unveiled any demonstrative associations [1]. This obvious paradox has led to increased targeting of diet balance patterns as a risk factor. In addition, three other risk factors that indicate a plausibly precancerous condition are sleep quality and psychosocial variables such as depression and anxiety [6]. However, relatively few studies have focused on connecting these lifestyle behaviors with HPV infection or HPV persistent infections.

HPV infection has been shown to be influenced by age, sexually transmitted diseases, and tumors. Existing studies have explored the link between HPV and four diseases including anemia [7], endocrine disease [8], metabolic disease [9, 10] and mental disorders [11]. They also cover HPV and three diseases in medical history – namely, reproductive tract infection [12, 13], tumors, and consanguinity tumors. Our study elucidates the relationship between lifestyle behavior factors and HPV infection.

A digital eHealth platform tool was developed on May 2020 to record age, lifestyle factors through the standard questionnaires and strategies, and disease status. Physical activity was monitored through the International Physical Activity Questionnaire (IPAQ) and could be beneficial for cancer control. Diet balance, sleep quality, depression and anxiety were recorded by the Diet Balance Index (DBI), Pittsburgh Sleep Quality Index (PSQI), Patient Depression Questionnaire-9 (PDQ-9), and Generalized Anxiety Disorder 7 (GAD-7) respectively. In addition, the digital eHealth platform provided information on HPV prevention courses via weekly live streaming while regularly providing advice for participants.

## Methods

### Participant Recruitment and Sample Collection

This study was approved by the ethics committee of the Institutional Review Board at Beijing Sports University (2021173H). From November 2021 to January 2022, study population were registered through the digital eHealth platform. Our team explained the study objectives and obtained signed informed consent from participants to collect questionnaires. Then, participants received a mailed package consisting of a nylon conical brush and graphic user instructions (SeqHPV; Beijing Genomics Institute, China). Participants were required to collect cervicovaginal mucus (CVM) samples by rotating brush and smear samples onto a Flinders Technology Associates (FTA) card. Participants were provided with a package and free express delivery to send their FTA cards to the laboratory. Personalized online guidance was offered if participants had any queries about sampling or delivery.

### Outcome Measures

Once the samples were returned to the laboratory, each FTA card was suspended in 5ml PreservCyt media in order for each self-sample to be sequenced for HPV (seqHPV) testing. These tests independently reported 16 serotypes in total, with 12 high-risk HPV serotypes (hrHPV) as HPV 16, 18, 31, 33, 35, 39, 45, 51, 52, 56, 58, 59 [14]; 2 intermediate risks as HPV 66 and 68 [15]; and 2 low risk HPV (lrHPV) as HPV 6 and 11 [16]. Participants received a test report online. Doctors also provided further advice to resolve any queries they had about their test.

Each lifestyle behavior had three categories of low, middle, and high risk, except for diet balance, which had an additional fourth category of extremely high risk. Physical activity was scored in three groups as low-risk (high physical activity), middle-risk (moderate physical activity), and high-risk (low physical activity) from IPAQ in Appendix 1 [17]. Diet balance was scored for Low-Bound Score (LBS) and High-Bound Score (HBS). LBS had four risk groups as low-risk -14 ∼ -1, middle-risk -29 ∼ -14, high-risk -43 ∼ -29, and extremely high-risk -72 ∼ -43. HBS also had four risk groups as low-risk 1 ∼ 9, middle-risk 10 ∼ 18, high-risk 19 ∼ 27, and extremely high-risk 27 ∼ 44 (DBI in Appendix 2). Sleep quality was scored in three groups as low-risk 0 ∼ 7, middle risk 7 ∼ 14, and high risk 14 ∼ 21 (PSQI in Appendix 3). Depression was scored in three groups as low-risk 0 ∼ 9, middle risk 9 ∼ 19, and high risk 19 ∼ 27 (PHQ-9, 0, 9, 19, 27 in in Appendix 4). Anxiety was scored in three groups as low-risk 0 ∼ 9, middle risk 9 ∼ 14, and high risk 14 ∼ 21 (GAD-7 in Appendix 5).

### Statistical Analysis

Baseline demographics were reported for two groups – one HPV-positive and the other HPV-negative. For the two groups, we summarized the continuous variables of age and lifestyle scores with both mean and standard deviation. Disease statuses were summarized as categorical variables with numbers and percentages. Differences were compared using the Wilcoxon rank sum test for continuous variables [18] and the Pearson χ^2^ test for categorical variables [19]. In addition, the statistical power was also calculated for lifestyle behavior factors.

In order to study the association between lifestyle behavior factors and HPV outcomes, we applied both univariable and multivariable logistic regression [20]. Regression was adjusted for age and disease status. We conducted further stratified analysis for different risk groups among lifestyle behavior factors. The reference group was low-risk for each lifestyle behavior factor.

To compare and assess the effect of HPV serotype, we differentiated the three risk groups for HPV infection as hrHPV (high-risk HPV), irHPV (intermediate-risk HPV) and lrHPV (low-risk HPV). We also enumerated the number of serotypes 1 ∼ 4 from participants. For one serotype (sole) and 2 ∼ 4 serotypes (multiple), physical activity and diet balance scores were tested by the Wilcoxon sum rank. All analysis was conducted utilizing R software (4.0.4) for the Macintosh Operating System (Mac OS).

## Results

### Participant Recruitment

In total, 495 participants were recruited via our digital eHealth tool with 149 HPV positive and 346 HPV negative women (Table 1). Patient features for HPV infection are elaborated in Table 1. The average age was 41.37 (SD: 7.86) for HPV positive and the average age for HPV negative was 41.56 (SD: 8.51), indicating no significant difference. Disease status was not statistically related to HPV infection. 28% of HPV positive participants and 24% of HPV negative participants had a history of reproductive tract infection. Among lifestyle behavior scores, there were statistically significant results for physical activity, diet balance, sleep quality, and depression. For physical activities (14.36 vs 12.83) and diet balance (25.44 vs 17.29), the higher scores indicate higher risk of HPV infection. They have a statistical power of 0.998 and over 0.999 respectively with *P-values* < 0.01. Conversely, another three lower scores denote higher risk on sleep scores (4.38 vs 4.94), depression (1.92 vs 2.27) and anxiety (2.75 vs 2.92). However, their statistical power reduced to 0.593 and 0.330 with *P-values* = 0.01.

**Table 1.** HPV Statistics for Age, Disease Status and Lifestyle Behavior.

| Risk factors | HPV positive (149) | HPV negative (346) | P-values<sup>1</sup> |
| --- | --- | --- | --- |
| Demography |  |  |  |
| Age | 41.37 ± 7.86 | 41.56 ± 8.51 | 0.29 |
| Disease status |  |  |  |
| Current Anemia | 21 (14.09%) | 67 (19.36%) | 0.16 |
| History of reproductive tract infection | 42 (28.19%) | 83 (23.99%) | 0.32 |
| Current endocrine disease | 12 (8.05%) | 23 (6.65%) | 0.58 |
| Current Metabolic disease | 4 (2.68%) | 12 (3.47%) | 0.65 |
| History of tumor disease | 10 (6.71%) | 20 (5.78%) | 0.69 |
| History of consanguinity tumor | 14 (9.40%) | 41 (11.85%) | 0.43 |
| Current Mental disorder | 2 (1.34%) | 3 (0.88%) | 0.63 |
| Lifestyle behavior scores (Questionnaires<sup>2</sup>) |  |  |  |
| Physical activity risk<sup>3</sup> (IPAQ) | 14.36 ± 1.95 | 12.83 ± 3.42 | <0.01### |
| Diet balance risk<sup>3</sup> (DBI-16) | 25.44 ± 7.33 | 17.29 ± 10.42 | <0.01### |
| Sleep quality scores<sup>3</sup> (PSQI) | 4.38 ± 3.28 | 4.94 ± 3.04 | 0.01## |
| Depression scores<sup>3</sup> (PHQ-9) | 1.92 ± 3.37 | 2.27 ± 2.98 | 0.01## |
| Anxiety scores (GAD-7) | 2.75 ± 5.53 | 2.92 ± 5.18 | 0.11 |

<sup>1</sup>: Statistical differences; ### denotes p-value < 0.01; ## denotes p-value = 0.01, # denotes 0.01 < p-value < 0.05; NA = Not available. <sup>2</sup>: IPAQ = International Physical Activity Questionnaires. DBI = Diet Balance Index, nutrition is negative score. PSQI = Pittsburgh Sleep Quality Index. PHQ-9 = Patient Depression Questionnaire-9. GAD 7 = Generalized Anxiety Disorder 7. <sup>3</sup>: Power of Physical activity = 0.998; Power of Diet Balance > 0.999; Power of Sleep = 0.593; Power of Depression = 0.330.

### Logistic Regression for Risk Factors

To evaluate the association of risk factors for HPV infection, both univariate and multivariate logistic regression were performed for one demography factor, seven disease status factors, and five lifestyle behavior factors (Table 2). For the left univariate logistic regression, physical activity significantly increased risk by 1.03 (95% CI: 1.02-1.05) and diet balance by 1.02 (95% CI: 1.01-1.02). For the subsequent right multivariate logistic regression, the increase was 1.20 (95% CI: 1.10-1.33) for physical activity and 1.09 (95% CI: 1.06-1.11) for diet balance, after considering the co-relationship with lifestyle behavior.

**Table 2.** Multivariable Logistic Regression for All Factors.

| Risk factors | Univariate logistic regression |  |  | Multivariable logistic regression |  |  |
| --- | --- | --- | --- | --- | --- | --- |
|  | Odds ratio | 95% CI | P-values | Odds ratio | 95% CI | P-values |
| Demography |  |  |  |  |  |  |
| Age | 1.00 | 1.00 - 1.00 | 0.792 | 1.01 | 0.99-1.04 | 0.320 |
| Disease status |  |  |  |  |  |  |
| Current Anemia | 1.08 | 0.97 - 1.20 | 0.160 | 1.48 | 0.87 - 2.59 | 0.159 |
| History of reproductive tract infection | 0.95 | 0.87 - 1.05 | 0.325 | 0.78 | 0.50 - 1.23 | 0.281 |
| Current endocrine disease | 0.96 | 0.82 - 1.12 | 0.577 | 0.82 | 0.40 - 1.79 | 0.613 |
| Current Metabolic disease | 1.05 | 0.84 - 1.33 | 0.652 | 1.61 | 0.53 - 6.08 | 0.432 |
| History of tumor disease | 0.97 | 0.82 - 1.14 | 0.691 | 0.78 | 0.35 - 1.84 | 0.555 |
| History of consanguinity tumor | 1.05 | 0.93 - 1.20 | 0.427 | 1.41 | 0.75 - 2.82 | 0.302 |
| Current Mental disorder | 0.90 | 0.60 - 1.36 | 0.629 | 0.76 | 0.12 - 5.93 | 0.768 |
| Lifestyle behavior |  |  |  |  |  |  |
| Physical activity (IPAQ) | 1.03 | 1.02 - 1.05 | < 0.001 | 1.20 | 1.10 - 1.33 | < 0.001 |
| Diet balance (DBI-16) | 1.02 | 1.01 - 1.02 | < 0.001 | 1.09 | 1.06 - 1.11 | < 0.001 |
| Sleep quality (PSQI) | 0.99 | 0.98 - 1.00 | 0.064 | 0.92 | 0.83 - 1.00 | 0.06 |
| Depression (PHQ-9) | 0.99 | 0.98 - 1.01 | 0.255 | 1.03 | 0.92 - 1.15 | 0.62 |
| Anxiety (GAD-7) | 1.00 | 0.99 - 1.01 | 0.751 | 0.99 | 0.94 - 1.05 | 0.76 |

### Lifestyle Behavior Factors

As expected, patients with high risk in physical activities were more likely to be infected with HPV in comparison to participants with low risk. This association was significant for p-value < 0.01. Patients with middle risk had a higher likelihood of being infected when compared with the low-level risk group (Table 3). Participants with middle, high, and extremely high-level diet imbalance had a higher possibility of being infected compared to participants with a low-risk diet balance. The middle level was significantly different. However, high-level and extremely high-level risk demonstrated no significant association between nutrition and HPV infection (Table 3). Another three lifestyle behavior factors were also tested: sleep quality risk, depression risk, and anxiety risk. However, they were not statistically significant. Although the probability of HPV infection increased with the risk of sleep quality and depression, the high-level risk of anxiety saw fewer HPV infections (Table 3).

**Table 3.** Univariable Logistic Regression: Risk of HPV Infection in Low, Middle and High-Level Risk Lifestyle Behavior Factors.

| Risk factors | HPV positive (149) | HPV negative (346) | Odds ratio | 95% CI | P-values |
| --- | --- | --- | --- | --- | --- |
| Physical activity risk |  |  |  |  |  |
| Low | 3 | 38 | 1.00 | NA | NA |
| Middle | 13 | 74 | 2.14 | 0.63-10.22 | 0.22 |
| High | 133 | 234 | 6.85 | 2.41-29.83 | <0.01<sup>##</sup> |
| Diet balance risk |  |  |  |  |  |
| Low | 15 | 163 | 1.00 | NA | NA |
| Middle | 129 | 147 | 0.11 | 0.06-0.18 | <0.01<sup>##</sup> |
| High | 4 | 28 | 0.63 | 0.21-2.41 | 0.46 |
| Extremely high | 1 | 8 | 0.66 | 0.11-17.52 | 0.78 |
| Sleep quality risk |  |  |  |  |  |
| Low | 126 | 284 | 1.00 | NA | NA |
| Middle | 22 | 59 | 1.18 | 0.70-2.06 | 0.52 |
| High | 1 | 3 | 1.22 | 0.14-35.31 | 0.80 |
| Depression risk |  |  |  |  |  |
| Low | 145 | 338 | 1.00 | NA | NA |
| Middle | 3 | 8 | 1.11 | 0.31-5.39 | 0.84 |
| High | 0 | 0 | NA | NA | NA |
| Anxiety risk |  |  |  |  |  |
| Low | 134 | 311 | 1.00 | NA | NA |
| Middle | 3 | 12 | 1.66 | 0.51-7.72 | 0.40 |
| High | 12 | 23 | 0.82 | 0.40-1.76 | 0.61 |

IPAQ = international physical activity questionnaires. DBI = diet balance index. PSQI = Pittsburgh sleep quality index. PHQ-9 = Patient Depression Questionnaire-9. GAD 7 = Generalized Anxiety Disorder 7. NA = Not available
Legend: CI, Confidence Interval; NA, Not available; statistically significant difference is marked <sup>##</sup> for P < 0.01.

### Physical Activity and Nutrition among HPV Serotypes

The prevalence of proportions involved 80.66% hrHPVs, 13.81% intermediate HPVs and 5.52% lrHPVs (Fig 1a). HPV 52 had the largest prevalence (19.89%) from the hrHPVs, followed by HPV16 (11.05%), then HPV 51 (9.39%). HPV 18 prevalence accounted for 4.42%. For intermediate HPV, HPV68 was more common than HPV66. However, the proportion was the same for lrHPVs, HPV 11 and HPV 6. More than 80% of HPV infections involved one serotype (Fig 1b). We then plotted a bar chart of two groups with mean and SD for physical activity (Fig 1c). A similar chart was plotted for diet balance and the significance was denoted as P-value < 0.05 (Fig 1d).

**Figure 1.**
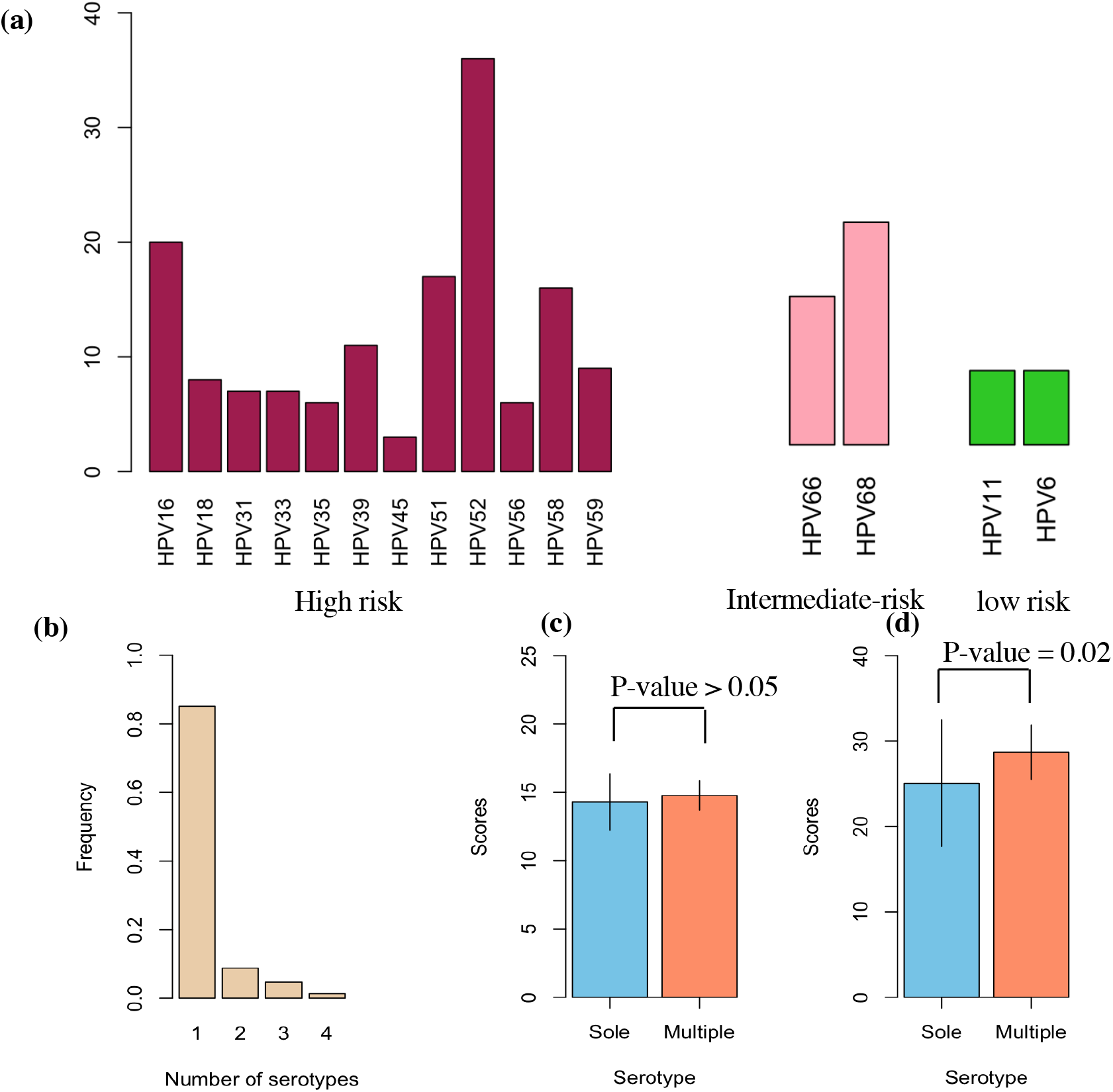
HPV Serotypes in Participants. (a)Each HPV serotype’s prevalence involved 12 high-risk HPV (hrHPV), 2 intermediate-risk HPV (irHPV), and 2 low-risk HPV (lrHPV). All identifiable serotypes were denoted at the x-axis, and their proportions as percentages were designated at the y-axis among all HPV-infected women. The left red columns indicate the prevalence of each serotype for the 12 hrHPV; the middle pink columns the prevalence of each serotype for the 2 irHPV; the right green columns the prevalence of each serotype for the 2 lrHPV. (b)This panel shows the number of serotypes identified for each patient infected with HPV. (c)Physical activity risk in two categories of data with sole and multiple serotypes in a bar chart. This plot displays the scores along with mean and SD without illustrating effect size. (d)Similar bar chart as (c), but with diet balance risk.

**Figure 2.**
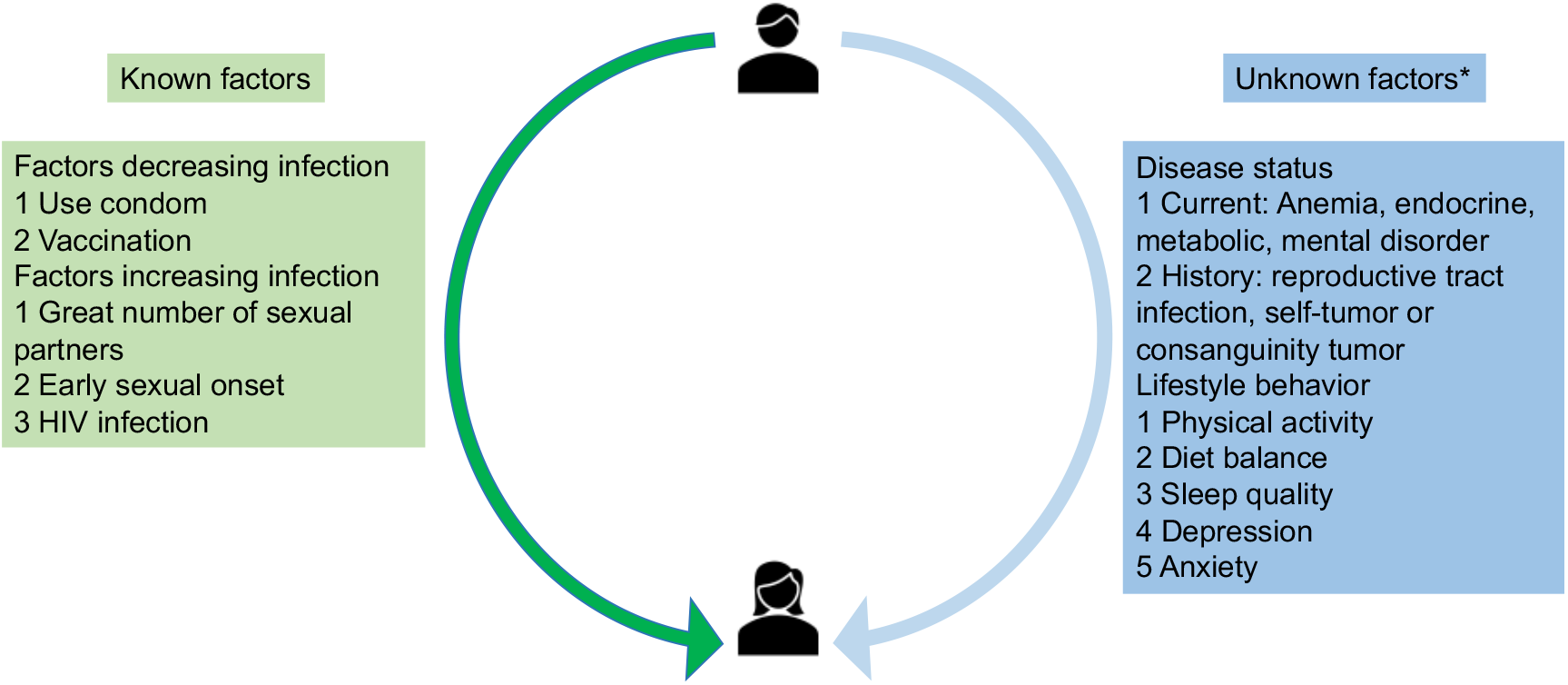
Transmission Cycle from Male to Female and Known/Unknown Factors Influencing Transmission Dynamics. HIV, human immunodeficiency virus. *Risk factors for the association direction was unclear before the study in this figure.

## Discussion

Most HPV risk factor studies focus on sexual factors or HIV infection in women. However, there are few cross-sectional studies that account for lifestyle risks or other gynecological infections. In our study, we surveyed 495 participants to investigate the connection between HPV infection and lifestyle behaviors as well as their gynecological disease history or current disease status. Apart from anxiety, four lifestyle factors appeared to demonstrate significant association with HPV infection: physical activity, diet balance, sleep quality, and depression. Meanwhile, current disease or disease history are not significantly correlated with HPV infection. After adjusting for age and disease status, multivariable logistic regression further showed significant association for physical activity and diet balance (P-values < 0.0001). In addition, there were 12 high-risk HPV (hrHPV) serotypes, more than the 2 intermediate-risk and 2 low-risk serotypes.

Our statistical analysis shed light on the most significant lifestyle behaviors. To enhance HPV infection through physical activity, stratification analysis reported the many benefits of low-risk physical activity. Potential reasons may include a decrease in monocyte engagement in the preinvasive tumor microenvironment (TME) causing a depletion in tumor-associated macrophages (TAMs) [21, 22]; or explained by the relation between HPV and human immune system like cell immunity such as CD4/CD8 T cell [23]. In other words, we recommend women infected with HPV to undertake higher levels of physical activity. Earlier studies have suggested two possible approaches for increasing physical activity [17]. The first involves vigorous intensive activity to achieve a minimum overall physical activity of no less than 1500 metabolic equivalents of task (MET) minutes for at least 3 days per week. The second involves achieving double the amount over the course of an entire week, or 3000 MET minutes every seven days, through any combination of vigorous or moderate intensity activities (which can even include walking).

To further enhance HPV infection through diet balance, stratification analysis showed that high-risk diet balance created greater difficulties for clearing HPV infection (OR: 3.27). This may be explained by difficulties in HPV infection being related to low levels of vitamin A or lycopene for high-risk diet balance [24, 25]; or also explained with the human immune system like cell immunity system [26]. Hence, we recommend two solutions for improving diet balance. Through our questionnaires, one way is to ensure appropriate consumption of dairy products and animal foods consisting of vitamin A through fruits (e.g., tomatoes) or vegetables in order to achieve a low-risk diet balance [27]. Beyond explanations from existing studies, the number of HPV serotypes is another potential justification. We found that diet balance scores were significantly increased among those with multiple HPV infections. Thus, the other solution implies that diet balance might be more effective for those with multiple HPV genotypes.

After adjusting for confounders, three lifestyle factors (sleep quality, depression, and anxiety) were shown to be insignificant in relation to HPV infection based on univariable and multivariable logistic regression. Furthermore, stratified analysis showed non-significant middle- and high-risk scores on sleep quality (*P-value*: 0.52 & 0.80); anxiety (*P-value*: 0.40 & 0.61); and high-risk depression (*P-value*: 0.84). Moreover, HPV infection does not appear to be associated with 7 gynecological diseases, whether current or in one’s history, as validated by univariable (*P-value*: 0.160 ∼ 0.652) and multivariable logistic regression (*P-value*: 0.159 ∼ 0.768) based on data collected from our eHealth platform.

Beyond lifestyle behaviors, our study also highlighted the benefits of communication via an eHealth platform. On our eHealth platform, participants actively responded to HPV advice. Through the platform, doctors provided weekly livestream training modules with advice about HPV self-sampling and cervical cancer prevention courses. During these sessions, participants were informed about the objective of our study and possible health benefits. In our previous pilot study in May 2020, our team established the eHealth platform and continues to serve HPV high-risk cohorts [28]. This regular training led to a high response rate from 2020 to 2021, as reflected by the growing audience size at our weekly livestream. Connecting with a wider audience has made it possible for us to plan further studies. Further ahead, we plan to enlarge the more widespread clinical variables considering the sexual factors. After that, the HPV infection model will be developed to predict the infection period through survival analysis.

### Strengths

In our study, one strength was the age range. The age range of HPV positive participants (41.37 ± 7.86) was comparable with HPV negative participants (41.56 ± 8.51). This age range is also the one in which women experience a high risk for HPV infection. Another strength was how the digital eHealth platform enabled participants to fill in their information electronically via their mobile phone. This substantially decreases the scope of human-made error. A third strength is how this study systematically considers the correlation between daily lifestyle behaviors and HPV infection after adjusting for the impact of age and current diseases or disease history.

### Weaknesses

There are three limitations to our study. First, other lifestyle behavior factors might affect common HPV infection risk, such as sexual behavior, smoking, or alcohol consumption. However, our study focused on determining HPV infection factors. Other studies have shown that HPV infection factors are different from infection factors. Second, our study locations consisted of more well-off areas such as Beijing and Shenzhen, where there is greater opportunity for abundant physical activity and dietary resources. A more comprehensive study would expand to other locations such as second-line cities or even the countryside in order to reduce geographical bias. To address this issue, our next formal study will focus on areas that encompass a broader scale, from less economically developed to more economically developed.

## Conclusions

This study suggests that both physical activity and diet balance are significantly beneficial lifestyle factors to reduce HPV infection. However, the three other factors of sleep quality, depression, and anxiety are required for the further evidence to prove any association. Finally, we recommend exercising regularly and regulating one’s diet balance might reduce the burden of HPV-related cancer such as cervical cancer.

## Supporting information

Supplemental survey 1

Supplemental survey 2

Supplemental survey 3

Supplemental survey 4

Supplemental survey 5

## Data Availability

All data produced in the present study are available upon reasonable request to the authors

## Contributors

Yantao Li and Zhongzhou Yang designed the project and carried out this study. Anli Wang found this project. Zhongzhou Yang and Mengping Liu analyzed the data and prepared the manuscript. Yantao Li collected the dataset and Anli Wang helped to obtain approval from the relevant ethics committees. Yantao Li, Mengping Liu and Anli Wang helped to further improve the quality of the manuscript.

## Acknowledgements

We acknowledge Tim Yung for English editing and the BGI team for developing the eHealth platform.

## References

1. Kerr J, Anderson C, Lippman SM. Physical activity, sedentary behaviour, diet, and cancer: an update and emerging new evidence. Lancet Oncol 2017; 18: e457–e471.

2. Bauman AE, Kamada M, Reis RS et al. An evidence-based assessment of the impact of the Olympic Games on population levels of physical activity. Lancet 2021; 398: 456–464.

3. Sallis JF, Bull F, Guthold R et al. Progress in physical activity over the Olympic quadrennium. Lancet 2016; 388: 1325–1336.

4. George SM, Ballard-Barbash R, Manson JE et al. Comparing indices of diet quality with chronic disease mortality risk in postmenopausal women in the Women’s Health Initiative Observational Study: evidence to inform national dietary guidance. Am J Epidemiol 2014; 180: 616–625.

5. Harmon BE, Boushey CJ, Shvetsov YB et al. Associations of key diet-quality indexes with mortality in the Multiethnic Cohort: the Dietary Patterns Methods Project. Am J Clin Nutr 2015; 101: 587–597.

6. Landau C, Novak AM, Ganz AB et al. Effect of Inquiry-Based Stress Reduction on Well-being and Views on Risk-Reducing Surgery Among Women With BRCA Variants in Israel: A Randomized Clinical Trial. JAMA Netw Open 2021; 4: e2139670.

7. Ferris RL, Saba NF, Gitlitz BJ et al. Effect of Adding Motolimod to Standard Combination Chemotherapy and Cetuximab Treatment of Patients With Squamous Cell Carcinoma of the Head and Neck: The Active8 Randomized Clinical Trial. JAMA Oncol 2018; 4: 1583–1588.

8. Yoon D, Lee JH, Lee H, Shin JY. Association between human papillomavirus vaccination and serious adverse events in South Korean adolescent girls: nationwide cohort study. BMJ 2021; 372: m4931.

9. Ilhan ZE, Laniewski P, Thomas N et al. Deciphering the complex interplay between microbiota, HPV, inflammation and cancer through cervicovaginal metabolic profiling. EBioMedicine 2019; 44: 675–690.

10. Lee J, Kim HS, Kim K et al. Metabolic syndrome and persistent cervical human papillomavirus infection. Gynecol Oncol 2021; 161: 559–564.

11. Sanders N, Miller B, Hernandez-Morales M, McEvoy J. Knowledge of HIV and HPV Among Women With Serious Mental Illness. Psychiatr Serv 2020; 71: 875.

12. Kriek JM, Jaumdally SZ, Masson L et al. Female genital tract inflammation, HIV co-infection and persistent mucosal Human Papillomavirus (HPV) infections. Virology 2016; 493: 247–254.

13. Spurgeon ME, Uberoi A, McGregor SM et al. A Novel In Vivo Infection Model To Study Papillomavirus-Mediated Disease of the Female Reproductive Tract. mBio 2019; 10.

14. Bouvard V, Baan R, Straif K et al. A review of human carcinogens--Part B: biological agents. Lancet Oncol 2009; 10: 321–322.

15. Cuzick J, Wheeler C. Need for expanded HPV genotyping for cervical screening. Papillomavirus Res 2016; 2: 112–115.

16. Dunne EF, Unger ER, Sternberg M et al. Prevalence of HPV infection among females in the United States. JAMA 2007; 297: 813–819.

17. Lear SA, Hu W, Rangarajan S et al. The effect of physical activity on mortality and cardiovascular disease in 130 000 people from 17 high-income, middle-income, and low-income countries: the PURE study. Lancet 2017; 390: 2643–2654.

18. Kumar S, Ooi CY, Werlin S et al. Risk Factors Associated With Pediatric Acute Recurrent and Chronic Pancreatitis: Lessons From INSPPIRE. JAMA Pediatr 2016; 170: 562–569.

19. Mahida JB, Asti L, Boss EF et al. Tracheostomy Placement in Children Younger Than 2 Years: 30-Day Outcomes Using the National Surgical Quality Improvement Program Pediatric. JAMA Otolaryngol Head Neck Surg 2016; 142: 241–246.

20. Zhou F, Yu T, Du R et al. Clinical course and risk factors for mortality of adult inpatients with COVID-19 in Wuhan, China: a retrospective cohort study. Lancet 2020; 395: 1054–1062.

21. Chang CW, Yang SF, Gordon CJ et al. Physical Activity of >/=7.5 MET-h/Week Is Significantly Associated with a Decreased Risk of Cervical Neoplasia. Healthcare (Basel) 2020; 8.

22. Goh J, Kirk EA, Lee SX, Ladiges WC. Exercise, physical activity and breast cancer: the role of tumor-associated macrophages. Exerc Immunol Rev 2012; 18: 158–176.

23. Niemiro GM, Coletta AM, Agha NH et al. Salutary effects of moderate but not high intensity aerobic exercise training on the frequency of peripheral T-cells associated with immunosenescence in older women at high risk of breast cancer: a randomized controlled trial. Immun Ageing 2022; 19: 17.

24. Kanetsky PA, Gammon MD, Mandelblatt J et al. Dietary intake and blood levels of lycopene: association with cervical dysplasia among non-Hispanic, black women. Nutr Cancer 1998; 31: 31–40.

25. Moore MA, Tajima K, Anh PH et al. Grand challenges in global health and the practical prevention program? Asian focus on cancer prevention in females of the developing world. Asian Pac J Cancer Prev 2003; 4: 153–165.

26. Perdigon G, Valdez JC, Rachid M. Antitumour activity of yogurt: study of possible immune mechanisms. J Dairy Res 1998; 65: 129–138.

27. Naresh A, Hagensee M, Myers L, Cameron J. Association of Diet Quality and Dietary Components with Clinical Resolution of HPV. Nutr Cancer 2021; 73: 2579–2588.

28. Zhongzhou Yang YZ, Araceli Stubbe-Espejel, Yumei Zhao, Mengping Liu, Jianjun Li, Yanping Zhao, Guoqing Tong, Na Liu, Andrew Hutchins, Songqing Lin, Yantao Li. Vaginal microbiota and personal risk factors associated with HPV status conversion – a new approach to reduce the risk of cervical cancer? PLoS One 2022.

