## Supplemental survey 1 for "The Lifestyle Factors of Physical Activity and Diet Balance associated with HPV Infection"

### Guidelines for Data Processing and Analysis of the International Physical Activity Questionnaire (IPAQ)

### – Short and Long Forms

#### November 2005

#### Contents

1. Introduction
2. Uses of IPAQ Instruments
3. Summary Characteristics of Short and Long Forms
4. Overview of Continuous and Categorical Analyses of IPAQ
5. Protocol for Short Form
6. Protocol for Long Form
7. Data Processing Rules
8. Summary Algorithms

Appendix 1. At A Glance IPAQ Scoring Protocol – Short Forms

Appendix 2. At A Glance IPAQ Scoring Protocol – Long Forms

#### 1. Introduction

This document describes recommended methods of scoring the data derived from the telephone / interview administered and self-administered IPAQ short and long form instruments. The methods outlined provide a revision to earlier scoring protocols for the IPAQ short form and provide for the first time a comparable scoring method for IPAQ long form. Latest versions of IPAQ instruments are available from www.ipaq.ki.se.

Although there are many different ways to analyse physical activity data, to date there is no formal consensus on a 'correct' method for defining or describing levels of physical activity based on self–report population surveys. The use of different scoring protocols makes it very difficult to compare within and between countries, even when the same instrument has been used. Use of these scoring methods will enhance the comparability between surveys, provided identical sampling and survey methods have been used.

#### 2. Uses of IPAQ Instruments

IPAQ short form is an instrument designed primarily for population surveillance of physical activity among adults. It has been developed and tested for use in adults (age range of 15-69 years) and until further development and testing is undertaken the use of IPAQ with older and younger age groups is not recommended.

IPAQ short and long forms are sometimes being used as an evaluation tool in intervention studies, but this was not the intended purpose of IPAQ. Users should carefully note the range of domains and types of activities included in IPAQ before using it in this context. Use as an outcome measure in small scale intervention studies is not recommended.

#### 3. Summary Characteristics of IPAQ Short and Long Forms

1. IPAQ assesses physical activity undertaken across a comprehensive set of domains including:
    a. leisure time physical activity
    b. domestic and gardening (yard) activities
    c. work-related physical activity
    d. transport-related physical activity;

2. The IPAQ **short** form asks about three specific types of activity undertaken in the four domains introduced above. The specific types of activity that are assessed are walking, moderate-intensity activities and vigorous-intensity activities.

3. The items in the **short** IPAQ form were structured to provide separate scores on walking, moderate-intensity and vigorous-intensity activity. Computation of the total score for the short form requires summation of the duration (in minutes) and frequency (days) of walking, moderate-intensity and vigorous-intensity activities. Domain specific estimates cannot be estimated.

4. The IPAQ **long** form asks details about the specific types of activities undertaken within each of the four domains. Examples include walking for transportation and moderate-intensity leisure-time activity.

5. The items in the **long** IPAQ form were structured to provide separate domain specific scores for walking, moderate-intensity and vigorous-intensity activity within each of the work, transportation, domestic chores and gardening (yard) and leisure-time domains. Computation of the total scores for the long form requires summation of the duration (in minutes) and frequency (days) for all the types of activities in all domains. Domain specific scores or activity specific sub-scores may be calculated. Domain specific scores require summation of the scores for walking, moderate-intensity and vigorous-intensity activities within the specific domain, whereas activity-specific scores require summation of the scores for the specific type of activity across domains.

#### 4. Overview of Continuous and Categorical Analyses of IPAQ

Both categorical and continuous indicators of physical activity are possible from both IPAQ forms. However, given the non-normal distribution of energy expenditure in many populations, it is suggested that the continuous indicator be presented as median minutes/week or median MET–minutes/week rather than means (such as mean minutes/week or mean MET-minutes/week).

##### 4.1 Continuous Variables

Data collected with IPAQ can be reported as a continuous measure. One measure of the volume of activity can be computed by weighting each type of activity by its energy requirements defined in METs to yield a score in MET–minutes. METs are multiples of the resting metabolic rate and a MET-minute is computed by multiplying the MET score of an activity by the minutes performed. MET-minute scores are equivalent to kilocalories for a 60 kilogram person. Kilocalories may be computed from MET-minutes using the following equation: MET-min x (weight in kilograms/60 kilograms). MET-minutes/day or MET-minutes/week can be presented although the latter is more frequently used and is thus suggested.

Details for the computation for summary variables from IPAQ short and long forms are detailed below. As there are no established thresholds for presenting MET-minutes, the IPAQ Research Committee propose that these data are reported as comparisons of median values and interquartile ranges for different populations.

##### 4.2 Categorical Variable: Rationale for Cut Point Values

There are three levels of physical activity proposed to classify populations:

1. Low
2. Moderate
3. High

The algorithms for the short and long forms are defined in more detail in Sections 5.3 and 6.3, respectively. Rules for data cleaning and processing prior to computing the algorithms appear in Section 7.

Regular participation is a key concept included in current public health guidelines for physical activity.[1] Therefore, both the total volume and the number of days/sessions are included in the IPAQ analysis algorithms.

The criteria for these levels have been set taking into account that IPAQ asks questions in all domains of daily life, resulting in higher median MET-minutes estimates than would have been estimated from leisure-time participation alone. The criteria for these three levels are shown below.

Given that measures such as IPAQ assess total physical activity in all domains, the "leisure time physical activity" based public health recommendation of 30 minutes on most days will be achieved by most adults in a population. Although widely accepted as a goal, in absolute terms 30 minutes of moderate-intensity activity is low and broadly equivalent to the background or basal levels of activity adult individuals would accumulate in a day. Therefore a new, higher cutpoint is needed to describe the levels of physical activity associated with health benefits for measures such as IPAQ, which report on a broad range of domains of physical activity.

**'High'**

This category was developed to describe higher levels of participation. Although it is known that greater health benefits are associated with increased levels of activity there is no consensus on the exact amount of activity for maximal benefit. In the absence of any established criteria, the IPAQ Research Committee proposes a measure which equates to approximately at least one hour per day or more, of at least moderate-intensity activity above the basal level of physical activity Considering that basal activity may be considered to be equivalent to approximately 5000 steps per day, it is proposed that "high active" category be considered as those who move at least 12,500 steps per day, or the equivalent in moderate and vigorous activities. This represents at least an hour more moderate-intensity activity over and above the basal level of activity, or half an hour of vigorous-intensity activity over and above basal levels daily. These calculations were based on emerging results of pedometers studies.[2]

This category provides a higher threshold of measures of total physical activity and is a useful mechanism to distinguish variation in population groups. Also it could be used to set population targets for health-enhancing physical activity when multi-domain instruments, such as IPAQ are used.

---

[1] Pate RR, Pratt M, Blair SN, Haskell WL , Macera CA, Bouchard C et al. Physical activity and public health. A recommendation from the Centers for Disease Control and Prevention and the American College of Sports Medicine. *Journal of American Medical Association* 1995; 273(5):402-7. and U.S. Department of Health and Human Services. *Physical Activity and Health: A Report of the Surgeon General.* Department of Health and Human Services, Centers for Disease Control and Prevention, National Center for Chronic Disease Prevention and Health Promotion, The Presidents' Council on Physical Fitness and Sports: Atlanta, GA:USA. 1996.

[2] Tudor-Locke C, Bassett DR Jr. How many steps/day are enough? Preliminary pedometer indices for public health. Sports Med. 2004;34(1):1-8.

**‘Moderate’**

This category is defined as doing some activity, more than the low active category. It is proposed that it is a level of activity equivalent to “half an hour of at least moderate-intensity PA on most days”, the former leisure time-based physical activity population health recommendation.

**‘Low’**

This category is simply defined as not meeting any of the criteria for either of the previous categories.

#### 5. Protocol for IPAQ Short Form

##### 5.1 Continuous Scores

Median values and interquartile ranges can be computed for walking (W), moderate-intensity activities (M), vigorous-intensity activities (V) and a combined total physical activity score. All continuous scores are expressed in MET-minutes/week as defined below.

##### 5.2 MET Values and Formula for Computation of MET-minutes/week

The selected MET values were derived from work undertaken during the IPAQ Reliability Study undertaken in 2000-2001[3]. Using the Ainsworth et al. Compendium (*Med Sci Sports Med* 2000) an average MET score was derived for each type of activity. For example; all types of walking were included and an average MET value for walking was created. The same procedure was undertaken for moderate-intensity activities and vigorous-intensity activities. The following values continue to be used for the analysis of IPAQ data: Walking = 3.3 METs, Moderate PA = 4.0 METs and Vigorous PA = 8.0 METs. Using these values, four continuous scores are defined:

Walking MET-minutes/week = 3.3 * walking minutes * walking days
Moderate MET-minutes/week = 4.0 * moderate-intensity activity minutes * moderate days
Vigorous MET-minutes/week = 8.0 * vigorous-intensity activity minutes * vigorous-intensity days
Total physical activity MET-minutes/week = sum of Walking + Moderate + Vigorous MET-minutes/week scores.

##### 5.3 Categorical Score

**Category 1 Low**

This is the lowest level of physical activity. Those individuals who not meet criteria for Categories 2 or 3 are considered to have a ‘low’ physical activity level.

[3] Craig CL,Marshall A , Sjostrom M et al. International Physical Activity Questionnaire: 12 country reliability and validity *Med Sci Sports Exerc* 2003;August

**Category 2 Moderate**

The pattern of activity to be classified as ‘moderate’ is either of the following criteria:

a) 3 or more days of vigorous-intensity activity of at least 20 minutes per day
   **OR**
b) 5 or more days of moderate-intensity activity and/or walking of at least 30 minutes per day
   **OR**
c) 5 or more days of any combination of walking, moderate-intensity or vigorous intensity activities achieving a minimum Total physical activity of at least 600 MET-minutes/week.

Individuals meeting at least one of the above criteria would be defined as accumulating a minimum level of activity and therefore be classified as ‘moderate’. See Section 7.5 for information about combining days across categories.

**Category 3 High**

A separate category labelled ‘high’ can be computed to describe higher levels of participation.
The two criteria for classification as ‘high’ are:

a) vigorous-intensity activity on at least 3 days achieving a minimum Total physical activity of at least 1500 MET-minutes/week
   **OR**
b) 7 or more days of any combination of walking, moderate-intensity or vigorous-intensity activities achieving a minimum Total physical activity of at least 3000 MET-minutes/week.

See Section 7.5 for information about combining days across categories.

#### 5.4 Sitting Question in IPAQ Short Form

The IPAQ sitting question is an additional indicator variable of time spent in sedentary activity and is not included as part of any summary score of physical activity. Data on sitting should be reported as median values and interquartile ranges. To-date there are few data on sedentary (sitting) behaviours and no well-accepted thresholds for data presented as categorical levels.

### 6. Protocol for IPAQ Long Form

The long form of IPAQ asks in detail about walking, moderate-intensity and vigorous-intensity physical activity in each of the four domains. Note: asking more detailed questions regarding physical activity within domains is likely to produce higher prevalence estimates than the more generic IPAQ short form.

#### 6.1 Continuous Score

Data collected with the IPAQ long form can be reported as a continuous measure and reported as median MET-minutes. Median values and interquartile ranges can be computed for walking (W), moderate-intensity activities (M), and vigorous-intensity activities (V) within each domain using the formulas below. Total scores may also be calculated for walking (W), moderate-intensity activities (M), and vigorous-intensity activities (V); for each domain (work, transport, domestic and garden, and leisure) and for an overall grand total.

#### 6.2 MET Values and Formula for Computation of MET-minutes

##### Work Domain

Walking MET-minutes/week at work = 3.3 * walking minutes * walking days at work

Moderate MET-minutes/week at work= 4.0 * moderate-intensity activity minutes * moderate-intensity days at work

Vigorous MET-minutes/week at work= 8.0 * vigorous-intensity activity minutes * vigorous-intensity days at work

Total Work MET-minutes/week =sum of Walking + Moderate + Vigorous MET-minutes/week scores at work.

##### Active Transportation Domain

Walking MET-minutes/week for transport = 3.3 * walking minutes * walking days for transportation

Cycle MET-minutes/week for transport= 6.0 * cycling minutes * cycle days for transportation

Total Transport MET-minutes/week = sum of Walking + Cycling MET-minutes/week scores for transportation.

##### Domestic and Garden [Yard Work] Domain

Vigorous MET-minutes/week yard chores= 5.5 * vigorous-intensity activity minutes * vigorous-intensity days doing yard work (**Note**: the MET value of 5.5 indicates that vigorous garden/yard work should be considered a moderate-intensity activity for scoring and computing total moderate intensity activities.)

Moderate MET-minutes/week yard chores= 4.0 * moderate-intensity activity minutes * moderate-intensity days doing yard work

Moderate MET-minutes/week inside chores= 3.0* moderate-intensity activity minutes * moderate-intensity days doing inside chores.

Total Domestic and Garden MET-minutes/week =sum of Vigorous yard + Moderate yard + Moderate inside chores MET-minutes/week scores.

##### Leisure-Time Domain

Walking MET-minutes/week leisure = 3.3 * walking minutes * walking days in leisure

Moderate MET-minutes/week leisure = 4.0 * moderate-intensity activity minutes * moderate-intensity days in leisure

Vigorous MET-minutes/week leisure = 8.0 * vigorous-intensity activity minutes * vigorous-intensity days in leisure

Total Leisure-Time MET-minutes/week = sum of Walking + Moderate + Vigorous MET-minutes/week scores in leisure.

#### Total Scores for all Walking, Moderate and Vigorous Physical Activities

Total Walking MET-minutes/week = Walking MET-minutes/week (at Work + for Transport + in Leisure)

Total Moderate MET-minutes/week total = Moderate MET-minutes/week (at Work + Yard chores + inside chores + in Leisure time) + Cycling Met-minutes/week for Transport + Vigorous Yard chores MET-minutes/week

Total Vigorous MET-minutes/week = Vigorous MET-minutes/week (at Work + in Leisure)

**Note:** Cycling MET value and Vigorous garden/yard work MET value fall within the coding range of moderate-intensity activities.

#### Total Physical Activity Scores

An overall total physical activity MET-minutes/week score can be computed as:

Total physical activity MET-minutes/week = sum of Total (Walking + Moderate + Vigorous) MET-minutes/week scores.

This is equivalent to computing:

Total physical activity MET-minutes/week = sum of Total Work + Total Transport + Total Domestic and Garden + Total Leisure-Time MET-minutes/week scores.

As there are no established thresholds for presenting MET-minutes, the IPAQ Research Committee proposes that these data are reported as comparisons of median values and interquartile ranges for different populations.

##### 6.3 Categorical Score

As noted earlier, regular participation is a key concept included in current public health guidelines for physical activity.[4] Therefore, both the total volume and the number of day/sessions are included in the IPAQ analysis algorithms. There are three levels of physical activity proposed to classify populations – 'low', 'moderate', and 'high'. The criteria for these levels are the same as for the IPAQ short [described earlier in Section 4.2]

**Category 1 Low**

This is the lowest level of physical activity. Those individuals who not meet criteria for Categories 2 or 3 are considered 'low'.

**Category 2 Moderate**

The pattern of activity to be classified as 'moderate' is either of the following criteria:

d) 3 or more days of vigorous-intensity activity of at least 20 minutes per day **OR**

e) 5 or more days of moderate-intensity activity and/or walking of at least 30 minutes per day

**OR**

[4] Pate RR, Pratt M, Blair SN, Haskell WL , Macera CA, Bouchard C et al. Physical activity and public health. A recommendation from the Centers for Disease Control and Prevention and the American College of Sports Medicine. *Journal of American Medical Association* 1995; 273(5):402-7. and U.S. Department of Health and Human Services. *Physical Activity and Health: A Report of the Surgeon General.* Department of Health and Human Services, Centers for Disease Control and Prevention, National Center for Chronic Disease Prevention and Health Promotion, The Presidents' Council on Physical Fitness and Sports: Atlanta, GA:USA. 1996.

f) 5 or more days of any combination of walking, moderate-intensity or vigorous-intensity activities achieving a minimum Total physical activity of at least 600 MET-minutes/week.

Individuals meeting at least one of the above criteria would be defined as accumulating a moderate level of activity. See Section 7.5 for information about combining days across categories.

**Category 3 High**

A separate category labelled 'high' can be computed to describe higher levels of participation.
The two criteria for classification as 'high' are:

a) vigorous-intensity activity on at least 3 days achieving a minimum Total physical activity of at least 1500 MET-minutes/week

**OR**

b) 7 or more days of any combination of walking, moderate-intensity or vigorous-intensity activities achieving a minimum Total physical activity of at least 3000 MET-minutes/week.

See Section 7.5 for information about combining days across categories.

##### 6.4 IPAQ Sitting Question IPAQ Long Form

The IPAQ sitting question is an additional indicator variable and is not included as part of any summary score of physical activity. To-date there are few data on sedentary (sitting) behaviours and no well-accepted thresholds for data presented as categorical levels. For the sitting question 'Minutes' is used as the indicator to reflect time spent in sitting rather than MET-minutes which would suggest an estimate of energy expenditure.

IPAQ long assesses an estimate of sitting on a typical weekday, weekend day and time spent sitting during travel (see transport domain questions).

##### Summary sitting variables include

Sitting Total Minutes/week = weekday sitting minutes* 5 weekdays + weekend day sitting minutes* 2 weekend days

Average Sitting Total Minutes/day = (weekday sitting minutes* 5 weekdays + weekend day sitting minutes* 2 weekend days) / 7

**Note:** The above calculation of 'Sitting Total' excludes time spent sitting during travel because the introduction in IPAQ long directs the responder to NOT include this component as it would have already been captured under the Transport section. If a summary sitting variable including time spent sitting for transport is required, it should be calculated by adding the time reported (travelling in a motor vehicle) under transport to the above formula. Care should be taken in reporting these alternate data to clearly distinguish the 'total sitting' variable from a 'total sitting – including transport' variable.

#### 7. Data Processing Rules

In addition to a standardized approach to computing categorical and continuous measures of physical activity, it is necessary to undertake standard methods for the cleaning and treatment of IPAQ datasets. The use of different approaches and rules would introduce variability and reduce the comparability of data.

There are no established rules for data cleaning and processing on physical activity. Thus, to allow more accurate comparisons across studies IPAQ Research Committee has established and recommends the following guidelines:

##### 7.1 Data Cleaning

I. Any responses to duration (time) provided in the hours and minutes response option should be converted from hours and minutes into minutes.
II. To ensure that responses in 'minutes' were not entered in the 'hours' column by mistake during self-completion or during data entry process, values of '15', '30', '45', '60' and '90' in the 'hours' column should be converted to '15', '30', '45', '60' and '90' minutes, respectively, in the minutes column.
III. In some cases duration (time) will be reported as weekly (not daily) e.g., VWHRS, VWMINS. These data should be converted into an average daily time by dividing by 7.
IV. If 'don't know' or 'refused ' or data are missing for time or days then that case is removed from analysis.

**Note**: Both the number of days *and* daily time are required for the creation of categorical and continuous summary variables

##### 7.2 Maximum Values for Excluding Outliers

This rule is to exclude data which are unreasonably high; these data are to be considered outliers and thus are excluded from analysis. All cases in which the sum total of all Walking, Moderate and Vigorous time variables is greater than 960 minutes (16 hours) should be excluded from the analysis. This assumes that on average an individual of 8 hours per day is spent sleeping.

The 'days' variables can take the range 0-7 days, or 8, 9 (don't know or refused); values greater than 9 should not be allowed and those cases excluded from analysis.

##### 7.3 Minimum Values for Duration of Activity

Only values of 10 or more minutes of activity should be included in the calculation of summary scores. The rationale being that the scientific evidence indicates that episodes or bouts of at least 10 minutes are required to achieve health benefits. Responses of less than 10 minutes [and their associated days] should be re-coded to 'zero'.

##### 7.4 Truncation of Data Rules

This rule attempts to normalize the distribution of levels of activity which are usually skewed in national or large population data sets.

In IPAQ short - it is recommended that all Walking, Moderate and Vigorous time variables exceeding ' 3 hours' or '180 minutes' are truncated (that is re-coded) to be equal to '180 minutes' in a new variable. This rule permits a maximum of 21 hours of activity in a week to be reported for each category (3 hours * 7 days).

In IPAQ long – the truncation process is more complicated, but to be consistent with the approach for IPAQ short requires that the variables total Walking, total Moderate-intensity and total Vigorous-intensity activity are calculated and then, for each of these summed behaviours, the total value should be truncated to 3 hours (180 minutes).

When analysing the data as categorical variable or presenting median and interquartile ranges of the MET-minute scores, the application of the truncation rule will not affect the results. This rule does have the important effect of preventing misclassification in the 'high' category. For example, an individual who reports walking for 10 minutes on 6 days and 12 hours of moderate activity on one day could be coded as 'high' because this pattern meets the '7 day" and "3000 MET-min" criteria for 'high'. However, this uncommon pattern of activity is unlikely to yield the health benefits that the 'high' category is intended to represent.

Although using median is recommended due to the skewed distribution of scores, if IPAQ data are analysed and presented as a continuous variable using mean values, the application of the truncation rule will produce slightly lower mean values than would otherwise be obtained.

##### 7.5 Calculating MET-minute/week Scores

Data processing rules 7.2, 7.3, and 7.4 deals first with excluding outlier data, then secondly, with recoding minimum values and then finally dealing with high values. These rules will ensure that highly active people remain classified as 'high', while decreasing the chances that less active individuals are misclassified and coded as 'high'.

Using the resulting variables, convert time and days to MET-minute/week scores [see above Sections 5.2 and 6.2; METS x days x daily time].

##### 7.6 Calculating Total Days for Presenting Categorical Data on Moderate and High Levels

Presenting IPAQ data using categorical variables requires the total number of 'days' on which all physical activity was undertaken to be assessed. This is difficult because frequency in 'days' is asked separately for walking, moderate-intensity and vigorous-intensity activities, thus allowing the total number of 'days' to range from a minimum of 0 to a maximum of 21'days' per week in IPAQ short and higher in IPAQ long. The IPAQ instrument does not record if different types of activity are undertaken on the same day.

In calculating 'moderately active', the primary requirement is to identify those individuals who undertake activity on at least '5 days'/week [see Sections 4.2 and 5.3]. Individuals who meet this criterion should be coded in a new variable called "*at least five days"* and this variable should be used to identify those meeting criterion b) at least 30 minutes of moderate-intensity activity and/or walking; and those meeting criterion c) any combination of walking, moderate-intensity or vigorous-intensity activities achieving a minimum of 600 MET-minutes/week.

Below are two examples showing this coding in practice:
i) an individual who reports '2 days of moderate-intensity' and '3 days of walking' should be coded as a value indicating "*at least five days"*;
ii) an individual reporting '2 days of vigorous-intensity', '2 days of moderate-intensity' and '2 days of walking should be coded as a value to indicate "*at least five days"* [even though the actual total is 6].

The original frequency of 'days' for each type of activity should remain in the data file for use in the other calculations.

The same approach as described above is used to calculate total days for computing the 'high' category. The primary requirement according to the stated criteria is to identify those individuals who undertake a combination of walking, moderate-intensity and or vigorous-intensity activity on at least 7 days/week [See section 4.2]. Individuals who meet this criterion should be coded as a value in a new variable to reflect *"at least 7 days*".

Below are two examples showing this coding in practice:
i) an individual who reports '4 days of moderate-intensity' and '3 days of walking' should be coded as the new variable *"at least 7 days*".
ii) an individual reporting '3 days of vigorous-intensity', '3 days moderate-intensity' and '3 days walking' should be coded as *"at least 7 days*" [even though the total adds to 9] .

#### 8. Summary algorithms

The algorithms in Appendix 1 and Appendix 2 to this document show how these rules work in an analysis plan, to develop the categories 1 [Low], 2 [Moderate], and 3 [High] levels of activity.

**IPAQ Research Committee**
**November 2005**

### APPENDIX 1

### At A Glance
#### IPAQ Scoring Protocol (Short Forms)

##### Continuous Score

Expressed as MET-min per week: MET level x minutes of activity/day x days per week

| **MET levels** | *Sample Calculation*<br>**MET-minutes/week for 30 min/day, 5 days** |
|---|---|
| Walking = 3.3 METs | 3.3*30*5 = 495 MET-minutes/week |
| Moderate Intensity = 4.0 METs | 4.0*30*5 = 600 MET-minutes/week |
| Vigorous Intensity = 8.0 METs | 8.0*30*5 = 1,200 MET-minutes/week |
| | TOTAL = 2,295 MET-minutes/week |

Total MET-minutes/week = Walk (METs*min*days) + Mod (METs*min*days) + Vig (METs*min*days)

##### Categorical Score- three levels of physical activity are proposed

**1. <u>Low</u>**

- No activity is reported **OR**
- Some activity is reported but not enough to meet Categories 2 or 3.

**2. <u>Moderate</u>**

Either of the following 3 criteria

- 3 or more days of vigorous activity of at least 20 minutes per day **OR**
- 5 or more days of moderate-intensity activity and/or walking of at least 30 minutes per day **OR**
- 5 or more days of any combination of walking, moderate-intensity or vigorous-intensity activities achieving a minimum of at least 600 MET-minutes/week.

**3. <u>High</u>**

Any one of the following 2 criteria

- Vigorous-intensity activity on at least 3 days and accumulating at least 1500 MET-minutes/week **OR**
- 7 or more days of any combination of walking, moderate- or vigorous-intensity activities accumulating at least 3000 MET-minutes/week

**Please review the full document "Guidelines for the data processing and analysis of the International Physical Activity Questionnaire" for more detailed description of IPAQ analysis and recommendations for data cleaning and processing [www.ipaq.ki.se].**

### APPENDIX 2

### At A Glance
#### IPAQ Scoring Protocol (Long Forms)

##### Continuous Score

Expressed as MET-minutes per week: MET level x minutes of activity/day x days per week

*Sample Calculation*

| **MET levels** | **MET-minutes/week for 30 min/day, 5 days** |
|---|---|
| Walking at work= 3.3 METs | 3.3*30*5 = 495 MET-minutes/week |
| Cycling for transportation= 6.0 METs | 6.0*30*5 = 900 MET-minutes/week |
| Moderate yard work= 4.0 METs | 4.0*30*5 = 600 MET-minutes/week |
| Vigorous intensity in leisure= 8.0 METs | 8.0*30*5 = 1,200 MET-minutes/week |
| | TOTAL = 3,195 MET-minutes/week |

<u>Domain Sub Scores</u>
Total MET-minutes/week at **work** = Walk (METs*min*days) + Mod (METs*min*days) + Vig (METs*min*days) at work

Total MET-minutes/week for **transportation** = Walk (METs*min*days) + Cycle (METs*min*days) for transportation

Total MET-minutes/week from **domestic and garden** = Vig (METs*min*days) yard work + Mod (METs*min*days) yard work + Mod (METs*min*days) inside chores

Total MET-minutes/week in **leisure-time** = Walk (METs*min*days) + Mod (METs*min*days) + Vig (METs*min*days) in leisure-time

<u>Walking, Moderate-Intensity and Vigorous-Intensity Sub Scores</u>

Total **Walking** MET-minutes/week = Walk MET-minutes/week (at Work + for Transport + in Leisure)

Total **Moderate** MET-minutes/week = Cycle MET-minutes/week for Transport + Mod MET-minutes/week (Work + Yard chores + Inside chores + Leisure) + Vigorous Yard chores MET-minutes

**Note:** The above is a total moderate activities only score. If you require a total of all moderate-intensity physical activities you would sum Total Walking and Total Moderate

Total **Vigorous** MET-minutes/week = Vig MET-minutes/week (at Work + in Leisure)

<u>Total Physical Activity Score</u>
**Total** Physical Activity MET-minutes/week = **Walking** MET-minutes/week + **Moderate** MET-minutes/week + Total **Vigorous** MET-minutes/week

Continued………..

**Also**

**Total** Physical Activity MET-minutes/week = Total MET-minutes/week (at Work + for Transport + in Chores + in Leisure)

#### Categorical Score- three levels of physical activity are proposed

1. **Low**

   No activity is reported **OR**
   a. Some activity is reported but not enough to meet Categories 2 or 3.

2. **Moderate**

   Either of the following 3 criteria
   a. 3 or more days of vigorous-intensity activity of at least 20 minutes per day **OR**
   b. 5 or more days of moderate-intensity activity and/or walking of at least 30 minutes per day **OR**
   c. 5 or more days of any combination of walking, moderate-intensity or vigorous-intensity activities achieving a minimum of at least 600 MET-min/week.

3. **High**

   Any one of the following 2 criteria
   - Vigorous-intensity activity on at least 3 days and accumulating at least 1500 MET-minutes/week **OR**
   - 7 or more days of any combination of walking, moderate- or vigorous- intensity activities accumulating at least 3000 MET-minutes/week

**Please review the full document "Guidelines for the data processing and analysis of the International Physical Activity Questionnaire" for more detailed description of IPAQ analysis and recommendations for data cleaning and processing [www.ipaq.ki.se].**
