## Supplemental survey 2 for "The Lifestyle Factors of Physical Activity and Diet Balance associated with HPV Infection"

### Table S1 DBI-16 components and standard for scoring[1]

| Component | Score | Subgroup | Score | Intake range by energy intake level | | | | | | | | | | |
|---|---|---|---|---|---|---|---|---|---|---|---|---|---|---|
| | | | | 1000 kcal | 1200 kcal | 1400 kcal | 1600 kcal | 1800 kcal | 2000 kcal | 2200 kcal | 2400 kcal | 2600 kcal | 2800 kcal | 3000 kcal |
| C1-Cereal | (-12)-12 | Cereal | (-12)-12 | 0g=-12<br>75-95g=0<br>>170g=12 | <15g=-12<br>90-110g=0<br>>185g=12 | 0g=-12<br>125-175g=0<br>>250g=12 | <10g=-12<br>175-225g=0<br>>390g=12 | <35g=-12<br>200-250g=0<br>>415g=12 | <5g=-12<br>225-275g=0<br>>495g=12 | <30g=-12<br>250-300g=0<br>>520g=12 | 0g=-12<br>275-325g=0<br>>600g=12 | <50g=-12<br>325-375g=0<br>>650g=12 | <75g=-12<br>350-400g=0<br>>675g=12 | <100g=-12<br>375-425g=0<br>>700g=12 |
| C2-Vegetable and fruit | (-12)-0 | Vegetable | (-6)-0 | ≥200g=0<br>160-199g=-1<br>Score decreased 1 with intake amount decreased 40g<br>0g=-6 | ≥250g=0<br>200-249g=-1<br>Score decreased 1 with intake amount decreased 50g<br>0g=-6 | ≥3000g=0<br>240-299g=-1<br>Score decreased 1 with intake amount decreased 60g<br>0g=-6 | | ≥400g=0<br>320-399g=-1<br>Score decreased 1 with intake amount decreased 80g<br>0g=-6 | ≥450g=0<br>360-449g=-1<br>Score decreased 1 with intake amount decreased 90g<br>0g=-6 | | ≥500g=0<br>400-499g=-1<br>Score decreased 1 with intake amount decreased 1000g<br>0g=-6 | | | ≥600g=0<br>480-599g=-1<br>Score decreased 1 with intake amount decreased 1200g<br>0g=-6 |
| | | Fruit | (-6)-0 | ⩾150g=0; 120-149g=-1<br>Score decreased 1 with intake amount decreased 30g<br>0g=-6 | | | ⩾2000g=0; 160-199g=-1<br>Score decreased 1 with intake amount decreased 40g<br>0g=-6 | | ⩾300g=0; 240-299g=-1<br>Score decreased 1 with intake amount decreased 60g<br>0g=-6 | | ⩾350g=0; 280-349g=-1<br>Score decreased 1 with intake amount decreased 70g<br>0g=-6 | | ⩾400g=0; 320-399g=-1<br>Score decreased 1 with intake amount decreased 80g<br>0g=-6 | |
| C3-Milk and dairy products Soybean and soybean products | (-12)-0 | Dairy | (-6)-0 | ≥500g=0<br>Score decreased 1 with intake amount decreased 100g<br>0g=-6 | | ≥350g=0<br>Score decreased 1 with intake amount decreased 70g<br>0g=-6 | ≥300g=0<br>Score decreased 1 with intake amount decreased 60g<br>0g=-6 | | | | | | | |
| | | Soybean | (-6)-0 | ⩾5g=0 | ⩾15g=0 | ⩾25g=0 | | | | | | | | |

| | | | | | | | | | | | | |
|---|---|---|---|---|---|---|---|---|---|---|---|---|
| | | | | Score decreased 1 with intake amount decreased 1g<br>0g=-6 | Score decreased 1 with intake amount decreased 3g<br>0g=-6 | | | | | Score decreased 1 with intake amount decreased 5g<br>0g=-6 | | |
| C4-Animal food | (-12)-8 | Red meat and products, Poultry and game | (-4)-4 | 0g=-3<br>1-5g=-2<br>6-10g=-1<br>11-20g=0<br>21-25g=1<br>26-30g=2<br>31-35g=3<br>>35g=4 | 0g=-4<br>1-5g=-3<br>6-10g=-2<br>11-15g=-1<br>16-35g=0<br>36-40g=1<br>41-45g=2<br>46-50g=3<br>>50g =4 | 0g=-4<br>1-10g=-3<br>11-20g=-2<br>21-30g=-1<br>31-50g=0<br>51-60g=1<br>61-70g=2<br>71-80g=-3<br>>80g =4 | | 0g=-4<br>1-15g=-3<br>16-30g=-2<br>31-45g=-1<br>46-55g=0<br>56-70g=1<br>71-85g=2<br>85-100g=3<br>>100g =4 | | 0g=-4<br>1-20g=-3<br>21-40g=-2<br>41-60g=-3<br>61-90g=0<br>91-110g=1<br>111-130g=2<br>131-150g=3<br>>150g=4 | 0g=-4<br>1-25g=-3<br>26-50g=-2<br>51-75g=-1<br>76-125g=0<br>126-150g=1<br>151-175g=2<br>176-200g=3<br>>200g=4 | |
| | | Fish and Shrimp | (-4)-0 | 0g=-4<br>1-4g=-3<br>5-9g=-2<br>10-14g=-1<br>⩾15g=0 | <5g=-4<br>5-9g=-3<br>10-14g=-2<br>15-19g=-1<br>⩾20g=0 | <10g=-4<br>10-19g=-3<br>20-29g=-2<br>30-39g=-1<br>⩾40g=0 | | <5g=-4<br>5-19g=-3<br>20-34g=-2<br>35-49g=-1<br>⩾50g=0 | | 0g=-4<br>1-24g=-3<br>25-49g=-2<br>50~74g=-1<br>⩾75g=0 | <25g=-4<br>25-49g=-3<br>50-74g=-2<br>75-99g=-1<br>⩾100g=0 | <50g=-4<br>50-74g=-3<br>75-99g=-2<br>100-124g=-1<br>⩾125g=0 |
| | | Egg | (-4)-4 | 0g=-4<br>1-5g=-3<br>6-10g=-2<br>11-15g=-1<br>16-25g=0<br>26-30g=1 | <5g=-4<br>6-10g=-3<br>11-15g=-2<br>16-20g=-1<br>21-30g=0<br>31-35g=1 | | 0g=-4<br>1-10g=-3<br>11-20g=-2<br>21-30g=-1<br>31-50g=0<br>51-60g=1 | | 0g=-4<br>1-15g=-3<br>16-30g=-2<br>31-45g=-1<br>46-55g=0<br>56-70g=1 | | | |

| | | | | | | | | | | |
|---|---|---|---|---|---|---|---|---|---|---|
| | | | | 31-35g=2<br>36-40g=3<br>>40g=4 | 36-40g=2<br>41-45g=3<br>>45g =4 | | 61-70g=2<br>71-80g=-3<br>>80g =4 | 71-85g=2<br>85-100g=3<br>>100g =4 | | |
| C5-Empty energy food | 0-12 | Cooking oil | 0-6 | ≤20g=0<br>21-25g=1<br>>45g=6 | ≤25g=0<br>26-30g=1<br>>50g=6 | | | | ≤30g=0<br>31-35g=1<br>>55g=6 | ≤35g=0<br>36-40g=1<br>>60g=6 |
| | | Alcoholic beverage | 0-6 | Male: ≤ 25g=0; 26-40g=1; score increased 1 with intake amount increased 15g; >100g=6<br>(25g alcohol=750ml beer or 250ml wine or 75g liquor 38°or 50g liquor > 38°)<br>Female: ≤15g=0; 16-25g=1; score increased 1 with intake amount increased 10g; >65g=6<br>(15g alcohol=450ml beer or 150ml wine or 50g liquor 38° or 30g liquor > 38°) | | | | | | |
| C6-Condiments | 0-12 | Addible sugar | 0-6 | ≤25g=0; 26g=1; score increased 1 with intake amount increased 5g; >50g=6 | | | | | | |
| | | Salt | 0-6 | <2g=0<br>2-3g=1<br>Score increased 1 with intake amount increased 2g<br>>12g=6 | <3g=0<br>3-4g=1<br>score increased 1 with intake amount increased 2g<br>>13g=6 | <4g=0<br>4-5g=1<br>Score increased 1 with intake amount increased 2g<br>14g=6 | <6g=0<br>6-7g=1<br>score increased 1 with intake amount increased 2g<br>>16g=6 | | | |
| C7-Diet variety | (-12)-0 | Diet variety | (-12)-0 | ≥12 kinds of food (soybean is 5g) =0; score decreased 1 with decreased 1 kinds of food | | | | | | |
| C8-Drinking water | (-12)-0 | Drinking water | (-12)-0 | ≥1200ml=0; score decreased 1 with intake amount decreased 100ml; <100ml=-12 | | | | | | |

[1]This table has been reproduced from He, Y., et al., Update of the Chinese diet balance index: DBI-16. Acta Nutrimenta Sinica, 2018. 40(06): p. 526-530.
