## Supplemental survey 3 for "The Lifestyle Factors of Physical Activity and Diet Balance associated with HPV Infection"

### The Pittsburgh Sleep Quality Index (PSQI)

**Name:** ________________________________________ **Date:** _______________________

**Instructions**: The following questions relate to your usual sleep habits during the past month only. Your answers should indicate the most accurate reply for the majority of days and nights in the past month. Please answer all questions.
During the past month,

**1**. When have you usually gone to bed? ___________________

**2**. How long (in minutes) has it taken you to fall asleep each night? ___________________

**3**. When have you usually gotten up in the morning? ___________________

**4**. How many hours of actual sleep do you get at night? (This may be different than the number of hours you spend in bed) __________________

Please check the appropriate blank below.

| **5**. During the past month, how often have you had trouble sleeping because you... | **Not during the past month (0)** | **Less than once a week (1)** | **Once or twice a week (2)** | **Three or More times a week (3)** |
|---|---|---|---|---|
| **a**. Cannot get to sleep within 30 minutes | **a.** _______ | __________ | __________ | __________ |
| **b**. Wake up in the middle of the night or early morning | **b.** _______ | __________ | __________ | __________ |
| **c**. Have to get up to use the bathroom | **c.** _______ | __________ | __________ | __________ |
| **d**. Cannot breathe comfortable | **d.** _______ | __________ | __________ | __________ |
| **e**. Cough or snore loudly | **e.** _______ | __________ | __________ | __________ |
| **f**. Feel too cold | **f.** _______ | __________ | __________ | __________ |
| **g**. Feel too hot | **g.** _______ | __________ | __________ | __________ |
| **h**. Have bad dreams | **h.** _______ | __________ | __________ | __________ |
| **i**. Have pain | **i.** ________ | __________ | __________ | __________ |
| **j**. Other reason(s), please describe, including how often you have had trouble sleeping because of this reason(s): | **j.** ________ | __________ | __________ | __________ |
| **6**. During the past month, how often have you taken medicine (prescribed or "over the counter") to help you sleep? | **6.** ________ | __________ | __________ | __________ |
| **7**. During the past month, how often have you had trouble staying awake while driving, eating meals, or engaging in social activity? | **7.** ________ | __________ | __________ | __________ |
| **8**. During the past month, how much of a problem has it been for you to keep up enthusiasm to get things done? | **8.** ________ | __________ | __________ | __________ |

| | **Very good (0)** | **Fairly good (1)** | **Fairly bad (2)** | **Very bad (3)** |
|---|---|---|---|---|
| **9**. During the past month, how would you rate your sleep quality overall? | **9.** ________ | __________ | __________ | __________ |

Physician Determined Global PSQI Score: **_________**
