## Supplemental survey 4 for "The Lifestyle Factors of Physical Activity and Diet Balance associated with HPV Infection"

#### PATIENT HEALTH QUESTIONNAIRE (PHQ-9)

ID #: ______________________ DATE: ______________

| Over the last *2 weeks,* how often have you been bothered by any of the following problems? *(use "✓" to indicate your answer)* | Not at all | Several days | More than half the days | Nearly every day |
|---|---|---|---|---|
| **1.** Little interest or pleasure in doing things | 0 | 1 | 2 | 3 |
| **2.** Feeling down, depressed, or hopeless | 0 | 1 | 2 | 3 |
| **3.** Trouble falling or staying asleep, or sleeping too much | 0 | 1 | 2 | 3 |
| **4.** Feeling tired or having little energy | 0 | 1 | 2 | 3 |
| **5.** Poor appetite or overeating | 0 | 1 | 2 | 3 |
| **6.** Feeling bad about yourself — or that you are a failure or have let yourself or your family down | 0 | 1 | 2 | 3 |
| **7.** Trouble concentrating on things, such as reading the newspaper or watching television | 0 | 1 | 2 | 3 |
| **8.** Moving or speaking so slowly that other people could have noticed. Or the opposite — being so figety or restless that you have been moving around a lot more than usual | 0 | 1 | 2 | 3 |
| **9.** Thoughts that you would be better off dead, or of hurting yourself | 0 | 1 | 2 | 3 |
| | add columns | ____ | + ____ | + ____ |

*(Healthcare professional: For interpretation of TOTAL, please refer to accompanying scoring card).* TOTAL: ______

| **10.** If you checked off *any problems,* how *difficult* have these problems made it for you to do your work, take care of things at home, or get along with other people? | |
|---|---|
| Not difficult at all | ______ |
| Somewhat difficult | ______ |
| Very difficult | ______ |
| Extremely difficult | ______ |

Copyright © 1999 Pfizer Inc. All rights reserved. Reproduced with permission. PRIME-MD© is a trademark of Pfizer Inc.

A2663B 10-04-2005

### PHQ-9 Patient Depression Questionnaire

#### For initial diagnosis:

1. Patient completes PHQ-9 Quick Depression Assessment.
2. If there are at least 4 ✓s in the shaded section (including Questions #1 and #2), consider a depressive disorder. Add score to determine severity.

##### *Consider Major Depressive Disorder*

- if there are at least 5 ✓s in the shaded section (one of which corresponds to Question #1 or #2)

##### *Consider Other Depressive Disorder*

- if there are 2-4 ✓s in the shaded section (one of which corresponds to Question #1 or #2)

**Note:** Since the questionnaire relies on patient self-report, all responses should be verified by the clinician, and a definitive diagnosis is made on clinical grounds taking into account how well the patient understood the questionnaire, as well as other relevant information from the patient.
Diagnoses of Major Depressive Disorder or Other Depressive Disorder also require impairment of social, occupational, or other important areas of functioning (Question #10) and ruling out normal bereavement, a history of a Manic Episode (Bipolar Disorder), and a physical disorder, medication, or other drug as the biological cause of the depressive symptoms.

**To monitor severity over time for newly diagnosed patients or patients in current treatment for depression:**

1. Patients may complete questionnaires at baseline and at regular intervals (eg, every 2 weeks) at home and bring them in at their next appointment for scoring or they may complete the questionnaire during each scheduled appointment.
2. Add up ✓s by column. For every ✓: Several days = 1 More than half the days = 2 Nearly every day = 3
3. Add together column scores to get a TOTAL score.
4. Refer to the accompanying **PHQ-9 Scoring Box** to interpret the TOTAL score.
5. Results may be included in patient files to assist you in setting up a treatment goal, determining degree of response, as well as guiding treatment intervention.

**Scoring: add up all checked boxes on PHQ-9**

**For every** ✓ Not at all = 0; Several days = 1;
More than half the days = 2; Nearly every day = 3

**Interpretation of Total Score**

| Total Score | Depression Severity |
|---|---|
| 1-4 | Minimal depression |
| 5-9 | Mild depression |
| 10-14 | Moderate depression |
| 15-19 | Moderately severe depression |
| 20-27 | Severe depression |

PHQ9 Copyright © Pfizer Inc. All rights reserved. Reproduced with permission. PRIME-MD ® is a trademark of Pfizer Inc.

A2662B 10-04-2005
