## Supplemental survey 5 for "The Lifestyle Factors of Physical Activity and Diet Balance associated with HPV Infection"

### Generalized Anxiety Disorder 7-Item (GAD-7) Scale

**Name:** Click here to enter text. **Date:** Click here to enter text.

| **Over the last 2 weeks, how often have you been bothered by the following problems?** | **Not At All** | **Several Days** | **Over Half the Days** | **Nearly Every Day** |
|---|---|---|---|---|
| **1. Feeling nervous, anxious, or on edge** | ☐ 0 | ☐ 1 | ☐ 2 | ☐ 3 |
| **2. Not being able to stop or control worrying** | ☐ 0 | ☐ 1 | ☐ 2 | ☐ 3 |
| **3. Worrying too much about different things** | ☐ 0 | ☐ 1 | ☐ 2 | ☐ 3 |
| **4. Trouble relaxing** | ☐ 0 | ☐ 1 | ☐ 2 | ☐ 3 |
| **5. Being so restless that it's hard to sit still** | ☐ 0 | ☐ 1 | ☐ 2 | ☐ 3 |
| **6. Becoming easily annoyed or irritable** | ☐ 0 | ☐ 1 | ☐ 2 | ☐ 3 |
| **7. Feeling afraid as if something awful might happen** | ☐ 0 | ☐ 1 | ☐ 2 | ☐ 3 |
| Add Scores for Each Column | ☐ | + ☐ | + ☐ | + ☐ |
| Total Score (Sum of Column Scores) | | | | |

**If any of the above problems were identified, how difficult have these made it for you to do your work, take care of things at home, or get along with other people?**

**☐ Not Difficult At All** **☐ Somewhat Difficult** **☐ Very Difficult** **☐ Extremely Difficult**

**Developed by Drs. Robert L. Spitzer, Janet B.W. Williams, Kurt Kroenke, and colleagues, with an education grant from Pfizer Inc. No permission required to reproduce, translate, display or distribute.**

### GAD-7 Important Notes and Scoring

The GAD-7 is based on the diagnostic criteria for GAD described in DSM-IV. *However, the GAD-7 is also sensitive to severity of symptoms of social phobia, post-traumatic stress disorder, and panic disorder.*

Please note: This questionnaire is designed for use by a health professional. Since the questionnaires rely on patient self-report, all responses should be verified by the clinician and a definitive diagnosis made on clinical grounds, taking into account how well the patient understood the questionnaire, as well as other relevant information from the patient (e.g., presence of DSM-IV GAD symptoms). A diagnosis of Generalized Anxiety Disorder should not be made based on GAD-7 scores alone.

***A score of 10 or greater indicates that further evaluation is required.***

Scoring Criteria: Total score (adding all the numbers) provides a possible score from 0-21.

GAD-7 Total Score Symptom Range

0-4 Minimal Anxiety
5-9 Mild Anxiety
10-14 Moderate Anxiety
15-21 Severe Anxiety

References:

Dear, B. F., Titov, N., McMillan, D., Anderson, T., Lorian, C., Robinson, E., & Sunderland, M. (2011). Psychometric comparison of the GAD-7 and PSWQ for measuring response during internet treatment for Generalised Anxiety Disorder. *Cognitive Behaviour Therapy, 40(3),* 216-227.

Löwe, B., Decker, O., Müller, S., Brähler, E., Schellberg, D., Herzog, W., & Herzberg, P. Y. (2008). Validation and standardization of the Generalized Anxiety Disorder Screener (GAD-7) in the general population. *Medical Care, 46,* 266-274.

Spitzer, R. L., Kroenke, K., Williams, J. B. W., & Löwe, B. (2006). A brief measure for assessing generalized anxiety disorder: The GAD-7. *Archives of Internal Medicine, 166*(10), 1092-1097. Swinson, R.P. (2006). The GAD-7 scale was accurate for diagnosing generalised anxiety disorder. *Evidence-Based Medicine, 11(6),* 184.
